# If long-term suppression is not possible, how do we minimize mortality for COVID-19 and other emerging infectious disease outbreaks?

**DOI:** 10.1101/2020.03.13.20034892

**Authors:** Andreas Handel, Joel C. Miller, Yang Ge, Isaac Chun-Hai Fung

## Abstract

As COVID-19 continues to spread, public health interventions are crucial to minimize its impact. The most desirable goal is to drive the pathogen quickly to extinction. This generally involves applying interventions as strongly as possible, which worked for SARS, but so far has failed for COVID-19. If fast eradication is not achievable, the next best goal is to delay the spread and minimize cases and burden on the health care system until suitable drugs or vaccines are available. This suppression approach also calls for strong interventions, potentially applied for a long time.

---

In the worst case, neither eradication nor suppression until vaccines become available are feasible. In such a circumstance, if infection induces immunity in individuals – which seems to be the case for coronavirus infections such as for SARS and MERS (1,2) and likely also COVID-19 (3) – the best option is to implement interventions such that new infections accrue slowly and do not overly strain the health care system, while allowing the number of susceptibles to drop until population (herd) immunity is achieved. At that point, interventions can be slowly relaxed without risking a resurgent epidemic. One goal in this situation is to implement control such that the total number of cases (the attack rate) is as low as possible. This is achieved by minimizing the overshoot, i.e., any excess infections that would deplete susceptibles beyond the population level immunity threshold. We discussed this idea previously (4–6).

Arguably, even more important than minimizing cases is minimizing deaths. For COVID-19, evidence so far suggests that mortality among children is low, while the elderly are at a much higher risk. This suggests that to reduce overall mortality, interventions should preferentially protect the elderly while allowing the minimum number of infections required to reach population immunity to occur in the least vulnerable groups. We illustrate how hypothetical interventions applied to different target groups affect overall mortality for COVID-19, using a simple simulation model (see Supplement for details). Three age groups, children, adults, and elderly, are considered, and interventions that reduce the risk of infection for each age group are implemented.

Scenario 1 in figure 1 shows a situation without interventions. Scenario 2 shows a situation where strong interventions are applied to protect all age groups. This reduces mortality the most while interventions are active, however, a subsequent outbreak occurs, and the total number infected and dead is similar to the no intervention scenario (7). Scenarios 3-5 show interventions that preferentially target children, adults, and the elderly, respectively. Each scenario leads to fewer infected and dead, despite weaker interventions compared to scenario 2. This is due to the prevention of a second large outbreak once interventions are removed. Importantly, while interventions 3-5 have a similar impact on the reduction of cases, the intervention targeting the elderly saves by far the most lives.

**Figure 1:**
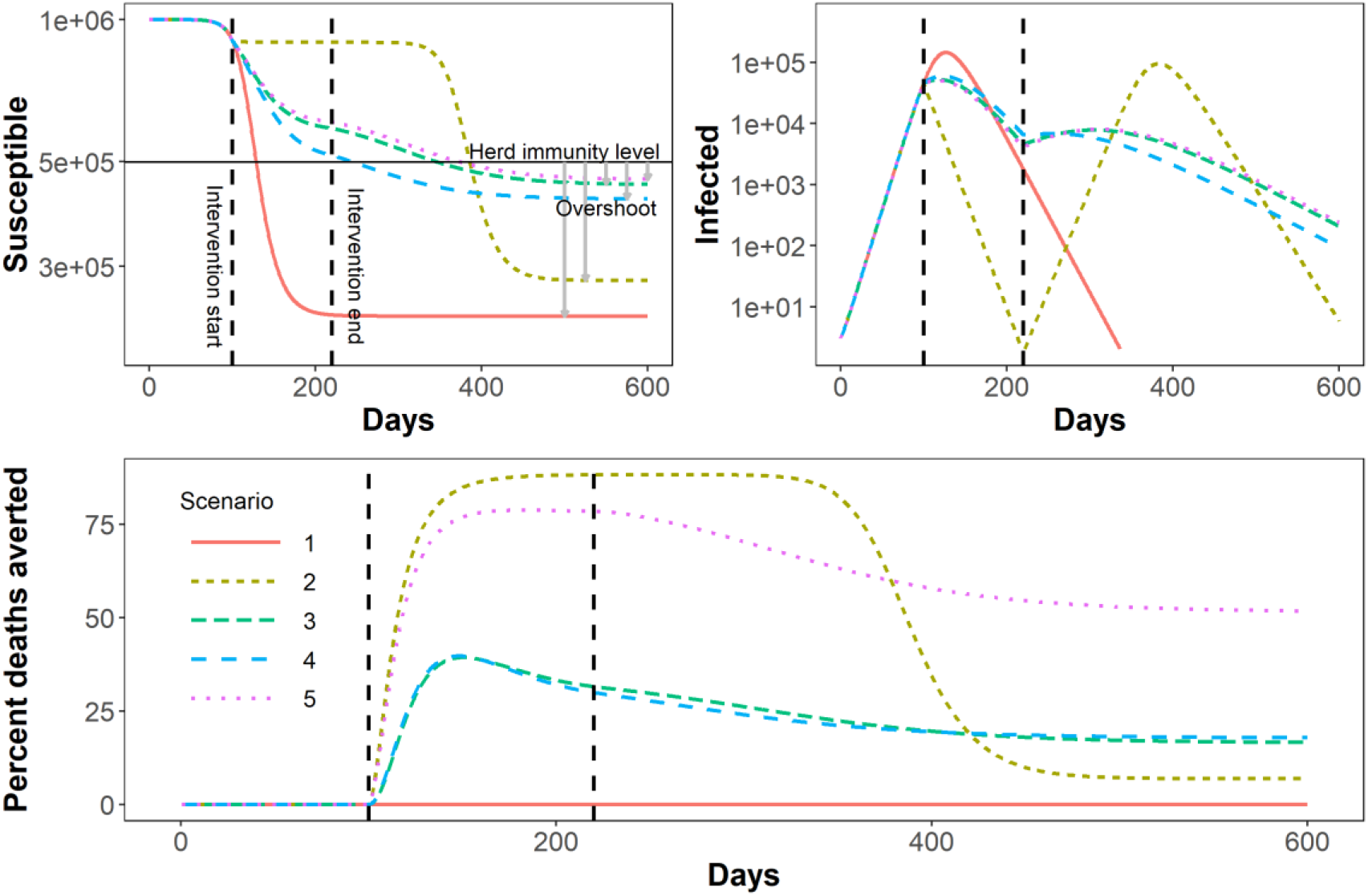
Susceptible, Infected, and percent deaths averted for different intervention scenarios. Scenario S1 shows a situation without any interventions, the maximum number of people get infected, the overshoot (gray arrow) is large, susceptibles drop far below the herd immunity level (solid horizontal line). In S2, strong interventions are applied to all groups, elderly are fully protected and children and adults have a reduction in infection rates of 90%. This is enough to drive down the outbreak while the interventions are active, thus initially reducing mortality the most. However, once the interventions are stopped, a consecutive outbreak occurs, leading to an overshoot and drop in susceptibles similar to the first scenario, and the overall number of averted deaths at the end of the outbreak is low. In S3-S5, the intervention prevents children, adults and the elderly respectively from getting infected. To ensure intervention strengths (effective reproductive numbers) are comparable between S3-S5, there is always some intervention applied to the adult group. Interventions start 100 days after outbreak start and last for 120 days (black vertical lines). See SM for details on the model and intervention implementation. Note that the model is very simple and only meant to illustrate our conceptual idea, it is not detailed enough to reflect any realistic intervention scenario.

This illustrates the importance of factoring in mortality among specific groups when deciding on the type of interventions to implement. Of course, it is worth emphasizing that there is still large uncertainty about the combined impact of our available interventions on the spread of COVID-19. The speed with which COVID-19 initially overwhelmed health systems in many countries, as well as the ability of several countries, to drive cases down successfully, suggests that the current best approach is to implement all feasible interventions against COVID-19 to suppress the disease. As the situation is monitored, it will become clear if we can accomplish elimination. If this is unlikely, and we are in a situation where interventions cannot be sustained long enough to prevent a resurgent epidemic, some interventions should be relaxed. Mitigation should then focus on allowing ‘controlled spread’ toward herd immunity in such a way that the most vulnerable populations are protected most, and the health care system remains functional. This might mean re-opening schools before re-starting other activities mainly frequented by adults or the elderly.

Since most interventions, including school-closures, have complex effects on contact patterns, it will be important to analyze the impact of different interventions using detailed simulation models and empirical studies. It will also be crucial to obtain solid estimates for the current number of infected and the number of individuals who have already been infected and recovered. This information is needed to estimate our location on the trajectory toward herd immunity and consequently how to adjust interventions. Decision-makers can then be adequately informed as they try to balance the need to relax interventions, keep transmission low, and reduce mortality.

## Data Availability

All code needed to reproduce all results is provided in the supplement.

